# ChronIQ-DRx2: restoring unwritten clinical decision logic unlocks personalized chronic disease management

**DOI:** 10.64898/2026.10.14.26365588

**Authors:** Yinzhi Jin, Yijing Li, Yuhang Shen, Jie Zhang, Zhouyue Wu, Yong Gao, Dan Su, Litao Li, Yanna Si, Yuwen Su, Dongning Yao, Xin Li, Weichao Ha, Feng Shao, Yingmei Lu, Danju Zhou, Qing Lan, Feng Han, Xun Chen

**Author notes:** Equal contribution (Yinzhi Jin, Yijing Li). Corresponding authors. **Correspondence:** Xun Chen.

## Abstract

Clinical records routinely document medical interventions but often omit the reasoning connecting patient-specific evidence to medical action. This limits the decision-relevant supervision available for training models for personalized care. Here we evaluate a data-augmentation approach that makes selected clinical decision relationships explicit in an 8-billion-parameter Qwen3 model, ChronIQ-DRx2. Using the same training cases and model architecture, expert-guided logic reconstruction was applied across four chronic diseases. On the held-out evaluation, the enhanced model had the highest plotted end-to-end task-completion rate among evaluated systems and higher plotted scores across diagnostic, medication and follow-up measures. The serious clinical error rates shown were 16.7% for the baseline and 12.5% for the enhanced model. In the human-AI evaluation, physician accuracy was 87.40% alone and 88.20% with AI assistance, while mean completion time was 0.959 and 0.780 min, respectively; medical-student accuracy was 62.71% and 74.80%, and mean completion time was 0.210 and 0.195 min. These results are descriptive point estimates; inferential comparisons are not reported here. Structured outputs may make recommendations easier to review, but rationale faithfulness, generalizability and effects on clinical safety require further evaluation.

## Introduction

Routine clinical records capture medical decisions yet consistently omit the underlying clinical decision logic. They document final diagnoses and prescribed drugs but discard the reasoning that links available evidence to specific conclusions or physiological constraints to altered treatments. This omission separates learning what clinicians did from when to do it. Consequently, the bottleneck in clinical modeling is not merely the volume of available data but the informational depth of each case.

Advanced medical AI such as AMIE have successfully extended automated capabilities into diagnostic dialogue and multivisit management. However, broad disease recognition and guideline concordance remain insufficient for complex clinical realities. True clinical proficiency requires nuanced judgments, such as recognizing when to request further investigations rather than prematurely concluding, prioritizing urgent referrals and understanding how specific organ functions dictate dosage adjustments. Standard models struggle with these individualized complexities because their training corpora lack these features. ^[5–10]^

Furthermore, automated performance does not eliminate the need for human clinical oversight. Structuring clinical decision relationships can provide targeted supervision during model training and produce outputs that are easier to inspect. However, generated rationales do not by themselves establish that a model’s stated reasoning faithfully represents its internal computation or improves clinical trust; these properties require direct evaluation. ^[11–15]^

Here we evaluate whether restoring selected clinical decision relationships is associated with changes in the diagnostic and prescribing performance of ChronIQ-DRx2, an 8-billion-parameter model. Without expanding the training case count or changing the model architecture, we combined algorithmic extraction with expert adjudication to structure decision relationships across stroke, type 2 diabetes, chronic hepatitis B and non-small-cell lung cancer. We compare the enhanced model with an unenhanced baseline across diagnosis, medication and follow-up tasks, and describe its performance in diagnostic-uncertainty, prescribing and human-AI evaluations. These comparisons characterize observed performance in the evaluated cohorts; further analyses are needed to establish statistical uncertainty, generalizability and clinical impact. ^[16–22]^

## Study workflow

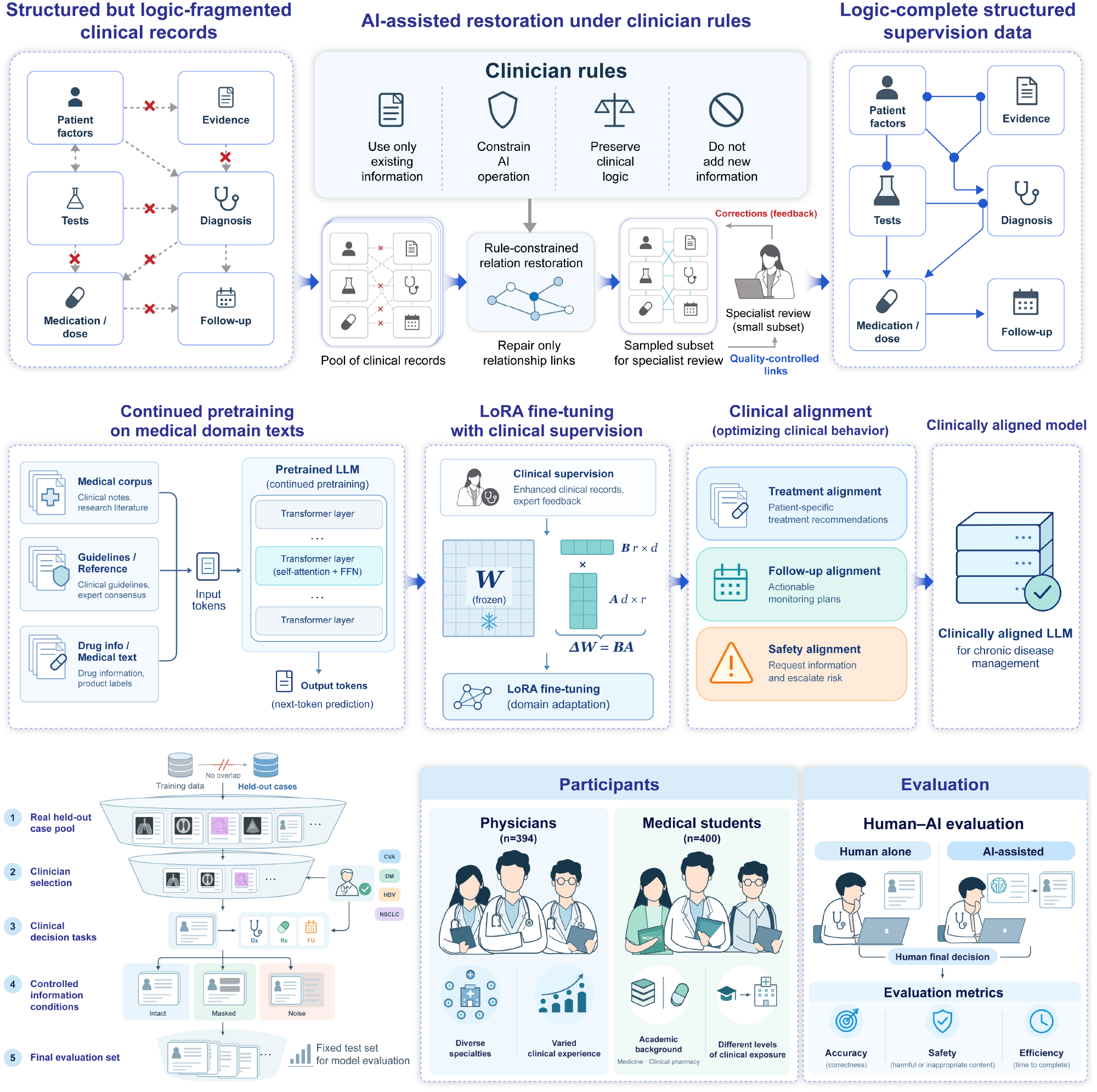

### Restoring clinical decision logic

Routine clinical records capture medical decisions but often discard the logic connecting clinical evidence to action. We quantified this deficit within a four-disease chronic care testbed spanning stroke, type 2 diabetes, chronic hepatitis B and non-small-cell lung cancer, comprising approximately 108,200 patients and 323,000 encounter-level training instances. Before restoration, diagnostic records were classified as missing or incomplete (14.2%), coarse (61.7%), refined (19.0%) or standardized (5.1%); medication records were classified as missing or insufficient (19.6%), drug-name-only (53.4%), standardized drug name (21.1%) or drug-disease-pathway linked (5.9%) (Fig. 1a). Follow-up records were most often absent (72.0%), with 22.0% containing general advice, 6.0% partially structured information and none fully structured. After restoration, logic-complete/standardized records accounted for 25.0% of diagnoses, 24.0% of medication records and 20.0% of follow-up records. Records also omitted critical links such as the pathological basis supporting a diagnosis, the findings defining a disease subtype and the physiological constraints dictating medication dosing. ^[1–4]^

**Fig. 1.**
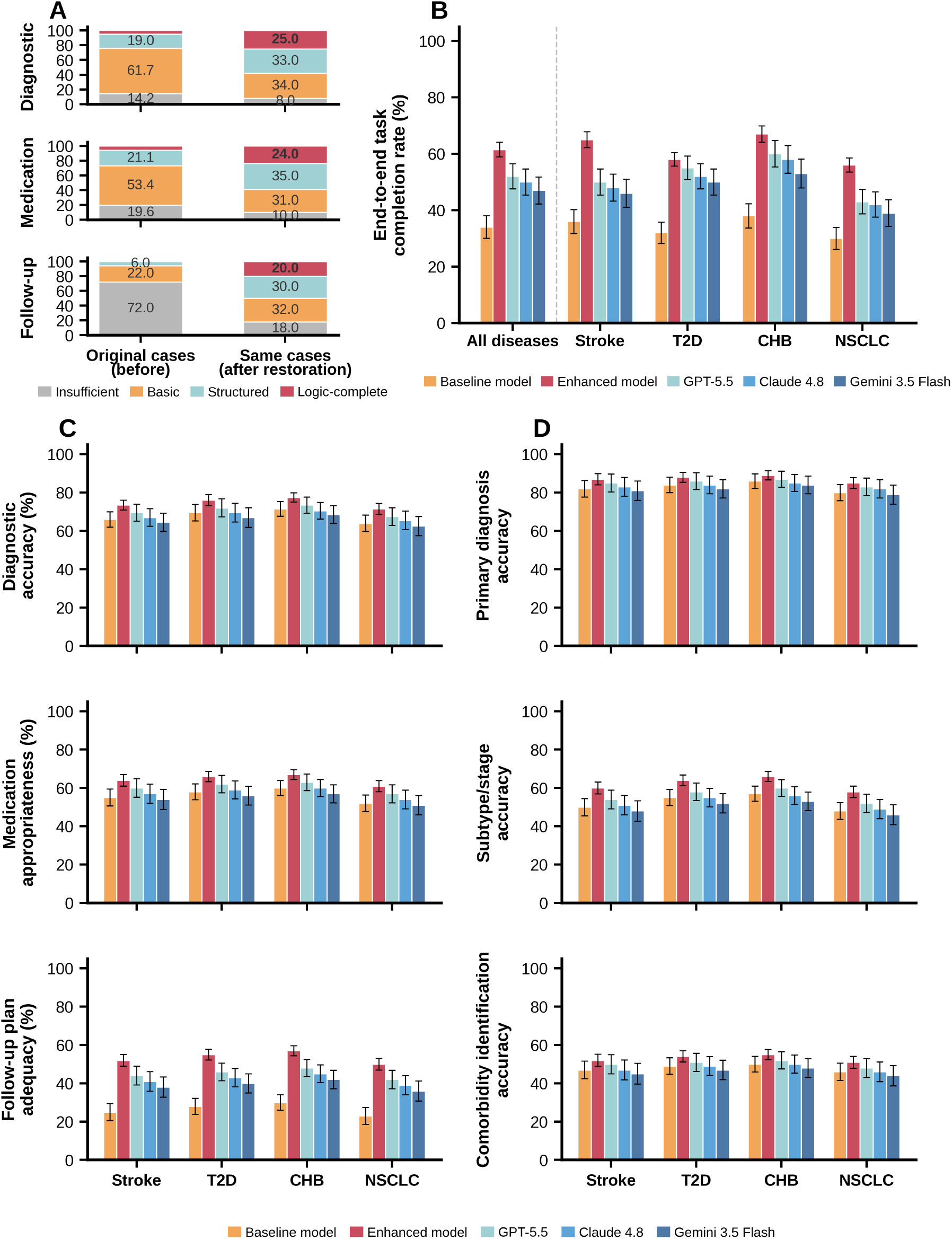
Expert enhancement of training corpora improves clinical model performance. a, Documentation-quality category distributions before and after restoration across diagnosis, medication and follow-up. b, End-to-end task completion across all diseases and disease-specific cohorts. c, Diagnostic accuracy, medication appropriateness and follow-up plan adequacy by disease. d, Primary diagnosis, subtype and comorbidity identification accuracy by disease. Error bars in b-d denote 95% confidence intervals.

Holding the 8B model architecture and training set size constant, training on this logic-restored corpus improved the diagnostic and prescribing accuracy of ChronIQ-DRx2 compared with the baseline model. End-to-end task completion was 61.5% for ChronIQ-DRx2 versus 34.0% for the baseline in the pooled cohort. The enhanced model also exceeded the baseline in stroke (65.0% versus 36.0%), type 2 diabetes (58.0% versus 32.0%), chronic hepatitis B (67.0% versus 38.0%) and NSCLC (56.0% versus 30.0%) (Fig. 1b).

Deconstructing this endpoint clarified the nature of the performance gains (Fig. 1c). Across the four diseases, diagnostic ability was 71.5–77.5% for the enhanced model versus 64.0–71.5% for the baseline; medication ability was 61.0–67.0% versus 52.0–60.0%; and follow-up plan adequacy was 50.0–57.0% versus 23.0–30.0%. The largest separation from the baseline was in follow-up planning, consistent with the sparse and fragmentary follow-up documentation in routine records.

Evaluation across diagnostic tasks showed that ChronIQ-DRx2 outperformed the baseline model in primary disease identification (85.0–89.0% versus 80.0–86.0%), clinical subtyping (58.0–66.0% versus 48.0–57.0%) and comorbidity identification (51.0–55.0% versus 46.0–50.0%) across the four disease cohorts (Fig. 1d). The separation was greatest for subtyping, whereas primary diagnosis and comorbidity showed smaller gains. These results distinguish broad disease recognition from more detailed clinical characterization.

Comorbidity identification showed a smaller separation between the enhanced and baseline models than clinical subtyping (Fig. 1d). Together with the domain-level results, this pattern indicates that gains varied by task, with the largest improvement in follow-up planning and a clear improvement in subtype classification.

### Diagnostic judgment under uncertainty

Clinical proficiency is most evident under diagnostic uncertainty. It requires recognizing which uncertainties matter and selecting investigations to resolve them. Across the four disease cohorts, appropriate diagnostic deferral was 86.7–90.0% for ChronIQ-DRx2, compared with 80.0–83.3% for the baseline and 73.3–80.0% for the generalist models. Out-of-scope recognition was 86.7% for ChronIQ-DRx2, 80.0–83.3% for the baseline and 70.0–80.0% for the generalist models. For targeted information requests, ChronIQ-DRx2 scored 83.3– 86.7%, versus 80.0–83.3% for the baseline and 73.3–80.0% for the generalist models (Fig. 2a). ^[23–26]^

**Fig. 2.**
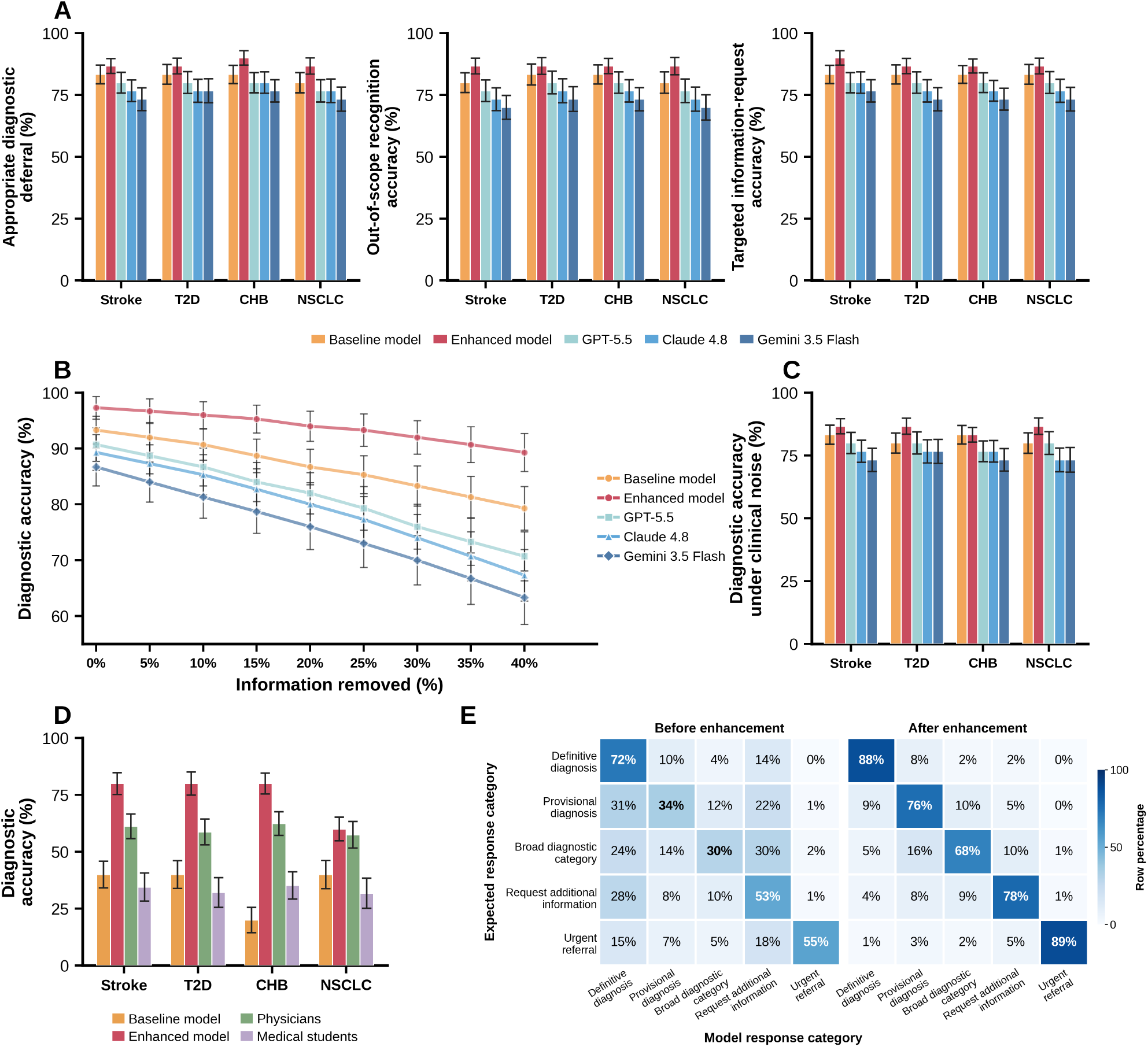
Diagnostic performance under uncertainty and perturbed information. a, Appropriate diagnostic deferral, out-of-scope recognition and targeted information-request accuracy across the four disease cohorts. b, Diagnostic accuracy as case information is progressively removed. c, Diagnostic accuracy after addition of noncontributory clinical information. d, Diagnostic accuracy compared with physician and medical-student reference groups. e, Row-normalized matrices comparing expected response categories (definitive diagnosis, provisional diagnosis, broad diagnostic category, request for additional information and urgent referral) with model response categories before and after enhancement. Error bars in a-d denote 95% confidence intervals.

Another dimension of clinical reasoning involves recognizing when missing information prevents a definitive diagnosis and when additional information is noncontributory. As information was progressively removed from 0% to 40%, diagnostic accuracy declined from 97.3% to 89.3% for ChronIQ-DRx2 and from 93.3% to 79.3% for the baseline; at 40% removal, the generalist models ranged from 63.3% to 70.7% (Fig. 2b). When non-contributory clinical information was added, ChronIQ-DRx2 accuracy was 83.3–86.7% across the four cohorts, compared with 80.0–83.3% for the baseline and 73.3–80.0% for the generalist comparators (Fig. 2c). These patterns are consistent with more robust interpretation of available evidence under the displayed perturbations.

We then compared model performance with physician and medical-student reference groups on a shared diagnostic completion task (Fig. 2d). Across stroke, type 2 diabetes, chronic hepatitis B and non-small-cell lung cancer, ChronIQ-DRx2 accuracy was 80%, 80%, 80% and 60%, respectively; corresponding baseline accuracy was 40%, 40%, 20% and 40%. Physician accuracy ranged from 57.5% to 62.4%, and medical-student accuracy from 31.8% to 35.2%. The enhanced model therefore had the highest plotted accuracy in three cohorts and was comparable to the physician group in the non-small-cell lung cancer cohort (60% versus 57.5%).

Analysis of the row-normalized response matrices shows how model responses aligned with the expected categories (Fig. 2e). Among cases assigned to the broad diagnostic category, responses classified as definitive diagnosis decreased from 24% before enhancement to 5% after enhancement. Among cases requiring a definitive diagnosis, responses classified as requests for additional information decreased from 14% to 2%. Among cases requiring urgent referral, responses classified as requests for additional information decreased from 18% to 5%. These displayed shifts indicate fewer premature definitive labels in broad-category cases and fewer information requests in cases requiring a definitive diagnosis or urgent referral.

### Individualized prescribing

Clinical guidelines provide a standard foundation for prescribing, yet their broad recommendations lack the individualized decision logic required for specific patients. Strict guideline concordance for ChronIQ-DRx2 was 85% for stroke, 82% for type 2 diabetes, 86% for chronic hepatitis B and 74% for non-small-cell lung cancer, compared with 78%, 72%, 82% and 68%, respectively, for the baseline. Generalist-model concordance ranged from 59% to 83% across diseases (Fig. 3a). ^[27–34]^

**Fig. 3.**
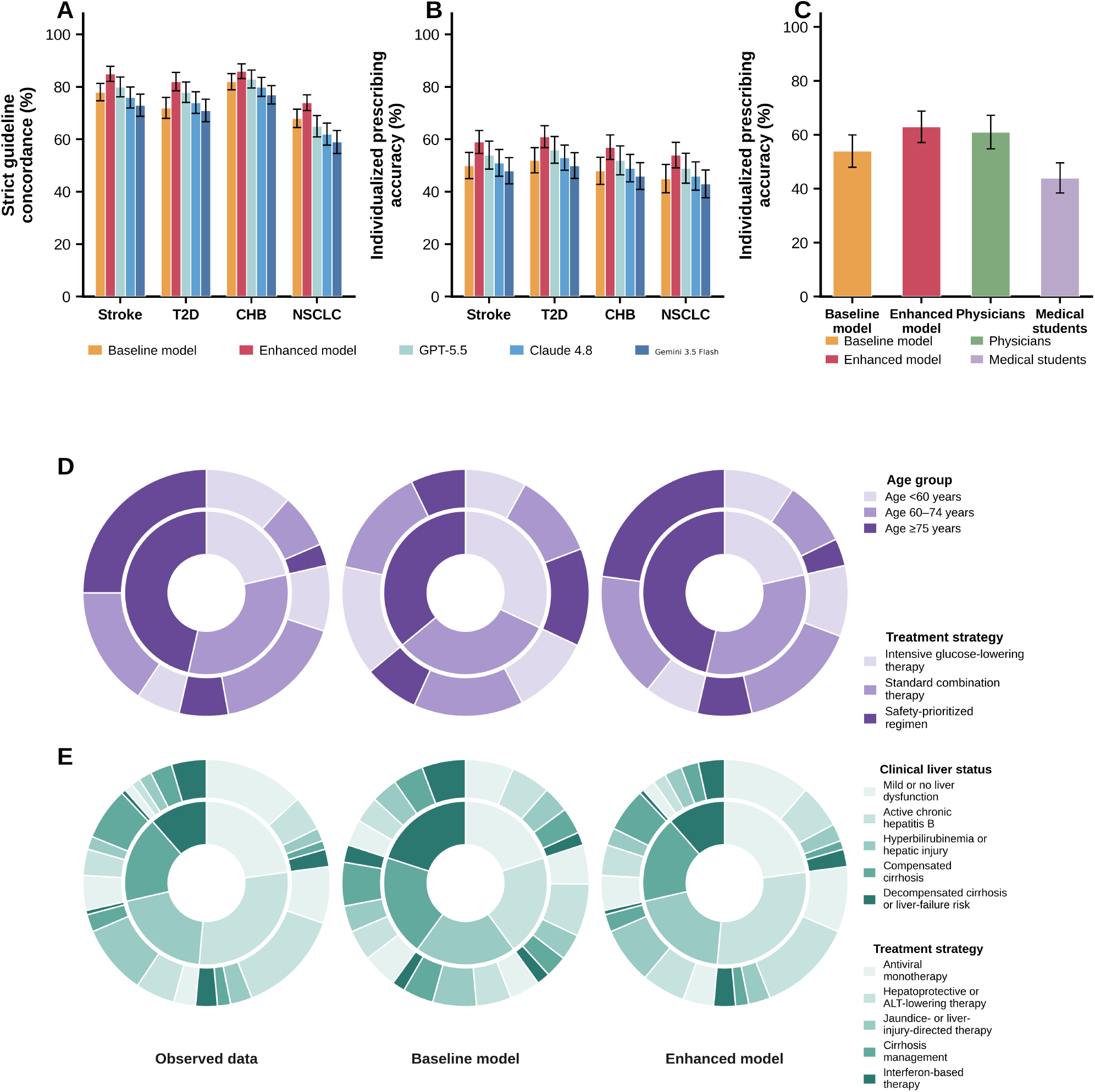
Guideline concordance, individualized prescribing and subgroup-specific treatment distributions. a, Strict guideline concordance across disease cohorts. b, Patient-specific prescribing accuracy across the four disease domains. c, Prescribing accuracy for the baseline and enhanced models and human reference groups. d, Diabetes treatment-strategy distributions across three age groups for observed data, the baseline model and the enhanced model. e, Hepatitis B treatment-strategy distributions across five clinical liver-status categories for observed data, the baseline model and the enhanced model. Error bars in a-c denote 95% confidence intervals.

The contribution of restored clinical decision logic was also apparent in individualized treatment regimens. Patient-specific prescribing accuracy for ChronIQ-DRx2 was 59% for stroke, 61% for type 2 diabetes, 57% for chronic hepatitis B and 54% for non-small-cell lung cancer. The corresponding baseline values were 50%, 52%, 48% and 45%, a 9-percentage-point difference in each cohort. Generalist-model accuracy ranged from 43% to 56% (Fig. 3b). These results show that the advantage was present in tasks requiring treatment choices to be adapted to individual patient circumstances.

In the human-reference comparison shown in Fig. 3c, individualized-prescribing accuracy was 63% for ChronIQ-DRx2, 54% for the baseline, 61% for physicians and 44% for medical students.

The subgroup panels present descriptive treatment-strategy distributions for observed data, the baseline model and the enhanced model. In the diabetes panel, the share of safety-prioritized treatment was 13.3%, 20.0% and 53.8% in the observed data for the three age groups (<60, 60–74 and ≥75 years), compared with 40.0%, 22.5% and 20.0% for the baseline and 16.7%, 22.2% and 49.2% for ChronIQ-DRx2 (Fig. 3d). The hepatitis B panel displays the distribution of five treatment strategies within each of five liver-status categories for the observed data and both models (Fig. 3e). These subgroup distributions are descriptive and are not presented as inferential comparisons.

### Structured clinical reasoning outputs

The model generated structured outputs that linked clinical findings with diagnostic considerations, medication choices and follow-up plans. Fig. 4a shows a representative stroke-prevention scenario in which these elements are presented as an organized sequence. The figure illustrates the output format; it is not a quantitative comparison of diagnostic accuracy. Although these outputs expose stated evidence and recommendations for review, this study did not evaluate whether the generated rationale faithfully reflects the model’s internal computation or enables clinicians to detect errors. ^[13–15,47,48]^

**Fig. 4.**
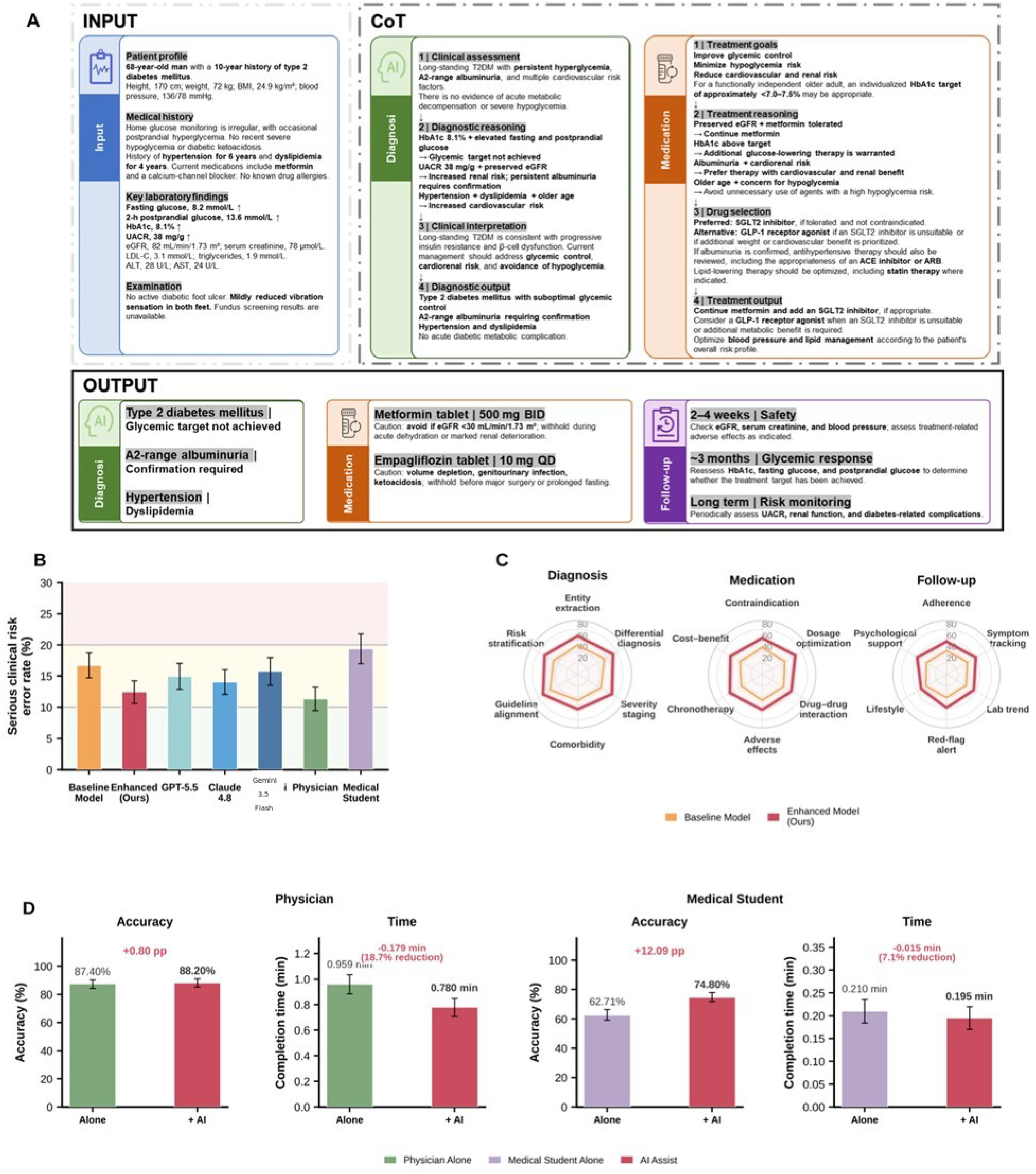
Auditable clinical decision logic and its evaluation in human-AI collaboration. a, Representative structured decision output linking diagnosis, medication and follow-up logic. b, Serious clinical error rates for the baseline and enhanced models, general-purpose comparators, physicians and medical students. c, Radar-chart comparison of baseline and enhanced model scores across diagnostic, medication and follow-up dimensions. d, Human-AI collaborative evaluation showing physician accuracy increasing from 87.40% to 88.20% with mean completion time decreasing from 0.959 to 0.780 min, and medical-student accuracy increasing from 62.71% to 74.80% with mean completion time decreasing from 0.210 to 0.195 min. Error bars in b and d denote 95% confidence intervals.

In Fig. 4b, the plotted serious clinical error rate was lower for ChronIQ-DRx2 than for the baseline and general-purpose comparators; the physician group had the lowest plotted rate, while the medical-student group had the highest. In the diagnostic, medication and follow-up dimensions shown in Fig. 4c, ChronIQ-DRx2 had higher plotted scores than the baseline across the displayed domains. These are descriptive comparisons; the figure alone does not establish statistical significance or improved clinical safety.

In the human-AI evaluation (Fig. 4d), medical-student accuracy was 62.71% without assistance and 74.80% with AI assistance (+12.09 percentage points); mean task-completion time was 0.210 and 0.195 min, respectively (7.1% lower with assistance). Physician accuracy was 87.40% without assistance and 88.20% with assistance (+0.80 percentage points); mean task-completion time was 0.959 and 0.780 min, respectively (18.7% lower with assistance). Participants completed different cases in the two conditions, so these values are condition-level descriptive comparisons, not same-case before-and-after changes. No inferential test results are presented here.

## Discussion

Routine clinical records preserve medical decisions but may omit the decision relationships that connect evidence with action. In this study, we applied a restoration pipeline without expanding the clinical case count or changing the 8-billion-parameter model architecture. The enhanced model had higher plotted performance than the baseline across several evaluated measures. These findings support further investigation of decision-logic restoration as a data-augmentation approach, but do not establish it as the primary bottleneck in clinical AI or demonstrate general clinical proficiency. ^[4,44–46]^

Rather than simply matching disease taxonomies to symptom profiles, ChronIQ-DRx2 learned to navigate clinical uncertainty. Appropriate diagnostic deferral ranged from 86.7% to 90.0%, compared with 80.0–83.3% for the baseline, and accuracy remained 89.3% when 40% of information was removed, compared with 79.3% for the baseline. Individualized-prescribing accuracy exceeded baseline accuracy by 9 percentage points in each of the four disease cohorts. These behaviors are consistent with clinical decision logic as conditional links between a patient state and a subsequent intervention. By augmenting the data with clinical decision logic, the model better represented when to investigate, when to intervene and when to refer.

The structured outputs present evidence, recommendations and follow-up considerations in a format that may be easier to review. However, we did not test rationale faithfulness, error-detection benefit, clinician trust or downstream patient safety, so these properties should not be inferred from the output format alone. In the human-AI evaluation, the plotted physician and student point estimates showed higher accuracy and shorter completion time with assistance. Because participants did not receive the same cases in both conditions and inferential results are not reported here, these differences should be interpreted descriptively. Future studies should prospectively assess generalizability, rationale faithfulness and the effects of assistance on clinical decisions and patient outcomes. ^[47–51]^

## Graphical highlight

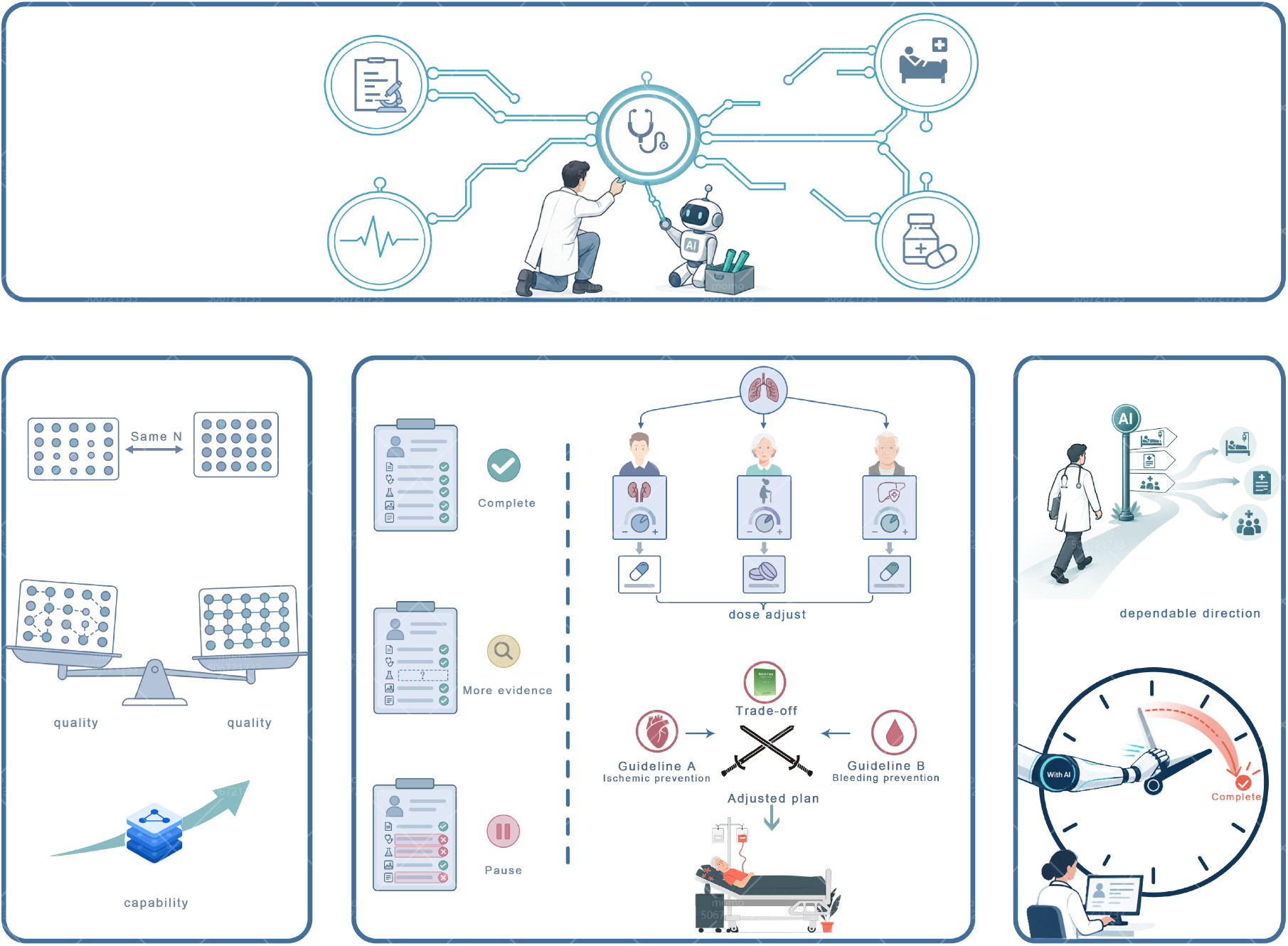

## Methods

### Study design

We developed and evaluated a post-training framework for chronic disease management in four clinical domains: stroke (CVA), type 2 diabetes mellitus (DM), chronic hepatitis B (HBV) and non-small-cell lung cancer (NSCLC). The study tested whether a combined clinical-information structuring and decision-logic restoration pipeline improved performance in diagnosis, individualized medication management and longitudinal follow-up. The baseline and enhanced models used the same model backbone, medical-domain continued-pretraining checkpoint, disease-specific LoRA-based supervised fine-tuning procedure and optimization settings. For each disease, the same encounter-level training instances were retained in both variants. The enhanced supervision corpus underwent the restoration pipeline described below, whereas the baseline corpus did not. The primary baseline-enhanced comparison therefore evaluates the combined restoration pipeline at fixed source patients, encounter identities, model backbone and optimization settings.

The source cohorts were initially indexed at the patient level, whereas model development and evaluation used the clinical encounter as the unit of analysis. Multiple encounters from the same patient could contribute to the training corpus. Data partitioning was performed at the patient level to prevent overlap between training and held-out evaluation sets. The held-out evaluation set contained 600 encounters, approximately 150 per disease, with one encounter per patient. The split was performed before model evaluation, and all model variants were evaluated on the same held-out cases.

### Real-world data and cohorts

Retrospective clinical records were obtained from a health-administration database aggregating data from multiple hospitals. The source data included demographics and basic clinical information, diagnostic records, laboratory measurements, imaging reports, medication records and narrative clinical documentation. Disease-specific cohorts were identified using combinations of diagnostic codes and text, investigations and medication information, followed by clinical confirmation according to predefined cohort rules. Cohort definitions combined diagnosis records, medication information, laboratory and imaging evidence, and clinical confirmation rules for each disease.

The source cohorts comprised approximately 38,200 patients with CVA, 49,300 with DM, 5,100 with HBV and 15,600 with NSCLC. Longitudinal records were reorganized around individual clinical encounters and decision points, yielding 98,600, 137,400, 29,800 and 57,200 encounter-level training instances, respectively. The difference between patient counts and encounter counts therefore reflects the change in analysis unit rather than the generation of additional patients.

The original database contained identifiable patient information. After extraction for research, analytic data were de-identified before case construction, augmentation, model training and evaluation. Identifiable source data were not used as model inputs.

### Case construction and preprocessing

Each encounter was organized around a clinical decision point. Model inputs comprised information available at that decision point, including chief complaint, history, laboratory data, imaging findings and relevant prior or current medication information. Targets comprised the diagnostic judgment, medication-management decision and follow-up plan. Information recorded only after the decision point was excluded from the model input to limit future-information leakage. Imaging inputs consisted of report text or structured imaging conclusions rather than raw images.

Clinical records underwent de-duplication, handling of missing or implausible values, unit harmonization and terminology normalization. Medication names and doses were standardized, and chronic-management regimens were separated from acute or short-term treatments that were not relevant to the target management setting. For example, treatments used during acute thrombectomy were not treated as chronic secondary-prevention therapy in the stroke cohort. Laboratory variables were standardized using disease-specific rules, including clinically relevant metabolic, hepatic, renal and virological measures. Disease-specific variable definitions were retained consistently across baseline and enhanced model development.

### Documentation-state assessment

To characterize the supervision available in routine records, documentation was assessed separately within the diagnosis, medication and follow-up domains. Records in each domain were assigned to one of four mutually exclusive documentation states: insufficient, basic, structured or logic-complete. These were descriptive categories used to quantify decision-relevant supervision quality and were not treated as a continuous clinical score.

Insufficient records lacked key facts or evidence required to support the corresponding clinical decision. Basic records contained recognizable diagnostic, medication or follow-up facts but remained incomplete, fragmented or limited to co-occurrence of clinical entities. Structured records contained sufficiently organized and identifiable clinical entities or fields, but the relationships between them remained largely implicit. Logic-complete records contained both the relevant clinical facts and explicit links between evidence or patient state, clinical judgment and action. Within each domain, the proportion in each state was calculated as the number of records assigned to that state divided by the total number of records in that domain.

The presence of clinical information was therefore distinguished from the presence of decision logic. For example, a record could contain both a diagnosis of ischemic stroke and aspirin as a medication without documenting that aspirin was selected for secondary prevention in the context of that diagnosis. Logic restoration converted such co-occurring facts into explicit decision-relevant relationships for supervision.

### Decision-logic restoration

The restoration pipeline combined rule-based processing, AI-assisted information structuring and physician-defined disease-specific logic. AI was used in two roles. First, it assisted extraction of clinical facts already present in unstructured text and converted them into structured fields. Second, after the clinical facts, physician-defined rules and decision relationships had been specified, AI was used to express these structured relationships as coherent clinical rationales for model supervision. AI-assisted processing was not permitted to overwrite original observed facts or independently redefine clinical decision labels. ^[35–40]^

The relationships targeted for restoration included evidence-to-primary-diagnosis links, evidence-to-subtype or stage links, missing-information-to-next-investigation links, patient-state-to-drug-selection links, physiological constraints on dose or treatment intensity, treatment-to-monitoring links and monitoring-result-to-treatment-adjustment links. The resulting supervision represented an explicit sequence from clinical evidence to judgment, action and longitudinal reassessment rather than a set of disconnected labels.

When decision-relevant evidence required by a predefined relation was absent from a training record, augmentation was constrained by observed case information and disease-specific clinical rules. Missing numerical evidence was represented as a clinically plausible range rather than as an exact observed measurement, and missing non-numerical evidence, including imaging-related information, was represented as a clinically consistent qualitative description. Original observations were preserved unchanged and augmented elements were treated as derived rather than observed information.

### Clinical rules and quality control

Clinical physicians defined a shared restoration framework and disease-specific rules using clinical guidelines, consensus recommendations and clinical experience. The rules specified the information required for diagnosis and subtyping, individualized medication selection and dosing, safety constraints, monitoring and follow-up, and conditions under which diagnosis should be deferred or escalation should occur.

Quality control was performed by stratified sampling across diseases and common clinical subtypes. Sampled restored records were independently reviewed by specialty physicians for factual consistency, clinical plausibility and adherence to the predefined rules. Reviewers could flag errors, document the reason for disagreement and return records for correction or reprocessing; disagreements were resolved by discussion and consensus. ^[41–43]^

### Continued pretraining and LoRA-SFT

Qwen3-8B-Instruct was used as the initial model backbone. The model underwent medical-domain continued pretraining on the publicly available Chinese-medical-dialogue corpus (xiaofengalg/Chinese-medical-dialogue; Apache License 2.0) using a causal language-modeling objective. This stage produced a shared medical-domain checkpoint used to initialize all subsequent disease-specific models.

For each disease, separate baseline and enhanced LoRA variants were trained independently from the same continued-pretraining checkpoint using identical implementation and optimization settings. LoRA rank was 64 and alpha was 128. Attention projections (q, k, v and o) and MLP projection layers were targeted. Batch size was 8 with gradient accumulation of 1, learning rate was 2 × 10™4, warm-up was 500 steps, the maximum number of training steps was 10,000, weight decay was 0.01 and optimization used 8-bit AdamW. Training stopped at the maximum number of steps. The same training configuration was used for baseline and enhanced variants within each disease.

### Comparator models and inference

General-purpose comparator models were evaluated on the same held-out cases using the same task instructions and output requirements within the comparator evaluation. The Gemini comparator was accessed through the third-party API relay endpoint ‘gemini_3_5_flash’. Calls were made programmatically through the same relay service on 5 July 2026 using de-identified case text. Each case was submitted once to each comparator; no retries, best-of-N selection or manual editing of model outputs was performed. Service-default inference settings were used when detailed decoding parameters were not exposed by the relay service.

### Held-out evaluation

All models were evaluated on the same 600 held-out encounters, with approximately 150 encounters per disease and no patient overlap with the training data. Reference answers were constructed using predefined clinical rules and guideline constraints and reviewed by clinical experts. Performance was assessed separately for diagnosis, medication management and follow-up. The overall chronic-management composite was the unweighted mean of the three component scores: (diagnosis score + medication score + follow-up score)/3. It was therefore a summary across management domains rather than a conjunctive endpoint requiring all three components to be simultaneously correct within one case. Reference adjudication followed the predefined clinical rule set before model-level comparisons were summarized.

Diagnostic analyses included primary disease identification, subtype or stage classification and comorbidity identification. Medication analyses assessed regimen appropriateness and patient-specific treatment selection. Follow-up analyses assessed adequacy of monitoring and longitudinal management plans. Failed or unparseable outputs were counted as incorrect.

### Missing-information and noise tests

Robustness to missing clinical information was evaluated by progressive masking of decision-relevant information rather than arbitrary text-token deletion. Physicians predefined disease- and task-specific checklists of clinically important information. For each case, missingness was calculated as the number of masked applicable decision-relevant items divided by the total number of applicable decision-relevant items. Masking was increased up to 40%. Only input evidence was masked; reference answers were unchanged, and all models were evaluated on identical masked case versions. Missingness analyses were applied identically across all evaluated models.

Noise robustness was evaluated by adding non-contributory clinical information sampled from real clinical records. Candidate symptoms, investigations or other details were reviewed to ensure that they should not alter the correct clinical conclusion for the target case, and the same noisy case version was presented to all models. Noise cases were generated before model comparison and were shared across models.

### Diagnostic uncertainty categories

To assess whether models adjusted diagnostic commitment to evidence sufficiency, each test condition was assigned to one of five clinically defined response categories: definitive diagnosis, diagnostic tendency, broad disease direction, insufficient information or urgent referral. Definitive diagnosis indicated sufficient evidence for a specific conclusion; diagnostic tendency indicated predominant support without full confirmation; broad disease direction indicated support only for a coarse disease category; insufficient information indicated that a definitive conclusion should be deferred pending additional information or investigations; and urgent referral indicated that predefined danger signals or management-boundary conditions required escalation.

Reference categories were generated from physician-defined information-sufficiency and decision-boundary rules rather than manually assigned case by case. A program applied these rules to the information available in each test condition so that the required response changed systematically when clinically important evidence was removed. The rule set produced one mutually exclusive reference category per test condition. Urgent referral and safety-boundary categories were given precedence when predefined danger signals were present.

### Individualized prescribing

Medication performance was evaluated against quality-controlled real-world prescribing references rather than treating every observed prescription as an absolute gold standard. Real-world prescriptions were screened using clinical rules and guideline boundaries to exclude clearly erroneous regimens, explicit contraindications and treatments inappropriate to the chronic-management context, such as acute-phase therapies when evaluating long-term stroke prevention.

Evaluation considered drug class and specific agent, dose or treatment intensity, organ-function constraints, contraindications, clinically relevant drug-drug interactions and appropriateness of combination regimens. Pre-specified subgroup analyses used clinically conventional stratification criteria identified from guideline and reference materials. Diabetes subgroup analyses used age-based strata, and hepatitis B subgroup analyses used clinically relevant liver-status strata.

### Critical clinical errors

A critical clinical error was defined as a model output with the potential to cause substantial patient harm, materially delay appropriate treatment or redirect subsequent management in a clinically important way. Physicians predefined the critical-error rules. Programmatic screening was first used to identify candidate errors, after which suspected critical errors were reviewed by clinicians; disagreements were resolved by discussion and consensus. A subset of non-flagged outputs was also reviewed for quality-control purposes.

Critical errors included clinically consequential missed or incorrect diagnoses, treatment recommendations involving explicit contraindications, unsafe doses or major drug-drug interactions, materially incorrect treatment direction, failure to escalate or refer when predefined danger criteria were present, and omission of monitoring or treatment-adjustment conditions essential for safety. Minor omissions, stylistic incompleteness or selection among multiple clinically acceptable alternatives were not automatically classified as critical errors.

### Physician ratings

Model outputs were independently assessed using a predefined multidimensional five-point anchored clinical rating rubric. The dimensions shown in Fig. 4c covered diagnosis (entity extraction, differential diagnosis, severity staging, comorbidity, guideline alignment and risk stratification), medication (contraindication handling, dosage optimization, drug-drug interaction checking, adverse effects, chronotherapy and cost-benefit) and follow-up (adherence, symptom tracking, laboratory trends, red-flag alerts, lifestyle and psychological support). Each dimension had explicit criteria for scores from 1 to 5, with higher scores indicating better performance. The complete scoring rubric is provided in the Supplementary Information.

For each disease cohort, the same fixed panel of five specialty physicians independently rated outputs from both the baseline and enhanced models for the same evaluation cases. Model identity was concealed during evaluation; raters viewed the case and generated response but not the model label. Ratings were paired by case and rater, with the same physicians assessing both model outputs for each case. Mean ratings across the five physicians were used for graphical presentation, whereas individual paired ratings were retained for statistical analysis. Cases and output order were organized to minimize recognizable model-order effects.

### Human-AI collaboration

The human-AI evaluation included 394 practicing physicians and 400 undergraduate students in medicine or clinical pharmacy. Physician participation comprised 115 participants in the CVA cohort, 96 in the DM cohort, 82 in the NSCLC cohort and 101 in the HBV cohort. One hundred students were recruited for each disease cohort. Physician seniority was recorded as a participant characteristic but was not used as a prespecified stratification factor. Participants were recruited from clinical and medical-education settings and completed the study using de-identified case materials.

Each participant completed ten disease-specific case tasks: five independently and five with AI assistance. Cases for both conditions were sampled from the same disease-specific pool. The same participant was not exposed to the same case under both conditions, although individual cases could be evaluated by different participants. In the AI-assisted condition, participants were shown the model’s structured reasoning together with its diagnostic, medication and follow-up recommendations. Task order and case allocation were balanced across independent and AI-assisted conditions.

For students, primary outcomes were diagnostic accuracy and critical clinical error rate. For practicing physicians, primary outcomes were diagnostic accuracy and task-completion time. Task-completion time was measured from case display to final answer submission.

## Supporting information

Supplementary Informatio

## Data Availability

The clinical data used in this study are not publicly available due to patient privacy and confidentiality restrictions. The trained model weights are also not publicly released to mitigate the risk of patient information being recovered from the model.

## Statistical analysis

Standalone model outcomes are summarized descriptively as proportions or scores, with 95% confidence intervals where indicated in the figure legends. No inferential comparisons are reported. The category distributions in Fig. 1a, row-normalized response matrices in Fig. 2e and subgroup treatment distributions in Fig. 3d,e are descriptive.

Human-AI study outcomes are summarized descriptively using the condition-specific point estimates shown in the figures. Participants completed different cases in the independent and AI-assisted conditions; therefore, comparisons are between study conditions and are not same-case paired before-and-after comparisons. Inferential tests are not reported.

## Ethics and data governance

This study was approved by the Ethics Committee of Nanjing Medical University (ethics review report No. 南医大伦审 (2025) 98 号; 6 June 2025). The study used de-identified chronic-disease clinical and medication records provided by the Jiangsu Health Information Center (Jiangsu TCM Information Center). Physicians and medical students participating in the human-AI evaluation volunteered to participate and provided informed consent before participation.

## AI-assisted writing

OpenAI Codex (GPT-5) was used to assist with manuscript writing and the organization and presentation of the results. The authors reviewed the AI-assisted content and take full responsibility for the final manuscript.

## Data availability

The clinical data used in this study contain sensitive patient information and are not publicly available. The trained model weights are also not publicly available because they were developed using sensitive clinical data. These restrictions are intended to protect patient privacy.

## Code availability

The code supporting this study is publicly available at https://github.com/jinyinzhi/ChronIQ_DRx2/. Clinical data and trained model weights are not publicly available because of privacy restrictions.

## Acknowledgements

We sincerely thank the physicians who participated in the human-AI evaluation and reviewed the disease-specific rules and restored records. We also thank the medical students who participated in the evaluation, the data-management staff who supported de-identification and secure case preparation, and the model-evaluation team for maintaining the comparator-model batch evaluation workflow and quality-control logs.

## Author Contributions

Yinzhi Jin, Yijing Li, Yuhang Shen, Jie Zhang and Zhouyue Wu contributed to data organization, figure preparation, model evaluation, literature integration and manuscript drafting. Yong Gao, Dan Su, Litao Li, Feng Shao, Yanna Si, Yuwen Su, Dongning Yao, Xin Li, Weichao Ha and Yingmei Lu contributed clinical-domain expertise, disease-specific rule review, case interpretation and evaluation design. Danju Zhou, Qing Lan, Feng Han and Xun Chen conceived and supervised the project, designed the clinical decision-logic restoration framework, interpreted results and revised the manuscript. All authors reviewed and approved the manuscript.

## Additional Information

Correspondence and requests for materials should be addressed to Xun Chen, the primary contact for this study. Supplementary information and reporting materials will be provided with the full submission package.

## References

1. Schiff, G. D. & Bates, D. W. Can electronic clinical documentation help prevent diagnostic errors? New England Journal of Medicine 362, 1066–1069 (2010). PMID 20335582.

2. Colicchio, T. K. & Cimino, J. J. Clinicians’ reasoning as reflected in electronic clinical note-entry and reading/retrieval: a systematic review and qualitative synthesis. Journal of the American Medical Informatics Association 26, 172–184 (2019). PMID 30576561.

3. Vuokko, R. et al. Impacts of structuring the electronic health record: results of a systematic literature review from the perspective of secondary use of patient data. International Journal of Medical Informatics 97, 293–303 (2017). PMID 27919387.

4. Tang, P. C. et al. Harnessing EHR data for health research. Nature Medicine 30, 1845–1853 (2024). PMID 38965433.

5. Elstein, A. S. & Schwarz, A. Clinical problem solving and diagnostic decision making: selective review of the cognitive literature. BMJ 324, 729–732 (2002). PMID 11909793.

6. Cook, D. A. et al. Management reasoning: beyond the diagnosis. JAMA 319, 2267–2268 (2018). PMID 29800012.

7. Moor, M. et al. Foundation models for generalist medical artificial intelligence. Nature 616, 259–265 (2023). PMID 37045921.

8. Tu, T. et al. Towards conversational diagnostic artificial intelligence. Nature 642, 103–113 (2025). PMID 40205050.

9. Liévin, V. et al. Towards conversational artificial intelligence for disease management. Nature 650, 100–110 (2026). PMID 42310463.

10. Chen, R. J. et al. LLM-assisted systematic review of large language models in clinical medicine. Nature Medicine 32, 1120–1132 (2026). PMID 41776077.

11. Hu, E. J. et al. LoRA: low-rank adaptation of large language models. ICLR (2022).

12. Lightman, H. et al. Let’s verify step by step. ICLR (2024).

13. Turpin, M. et al. Language models don’t always say what they think: unfaithful explanations in chain-of-thought prompting. NeurIPS (2023).

14. Lanham, T. et al. Measuring faithfulness in chain-of-thought reasoning. arXiv (2023).

15. Rosenbacke, R. et al. How explainable artificial intelligence can increase or decrease clinicians’ trust in AI applications in health care: systematic review. JMIR AI 3, e53207 (2024). PMID 39476365.

16. Langenberg, C., Hingorani, A. D. & Whitty, C. J. M. Biological and functional multimorbidity: from mechanisms to management. Nature Medicine 29, 1649–1657 (2023). PMID 37464031.

17. Barnett, K. et al. Epidemiology of multimorbidity and implications for health care, research, and medical education: a cross-sectional study. Lancet 380, 37–43 (2012). PMID 22579043.

18. Tobias, D. K. et al. Second international consensus report on gaps and opportunities for the clinical translation of precision diabetes medicine. Nature Medicine 29, 2438–2457 (2023). PMID 37794253.

19. GBD 2021 Nervous System Disorders Collaborators. Global, regional, and national burden of disorders affecting the nervous system, 1990-2021: a systematic analysis for the Global Burden of Disease Study 2021. Lancet Neurology 23, 344–381 (2024). PMID 38493795.

20. GBD 2021 Diabetes Collaborators. Global, regional, and national burden of diabetes from 1990 to 2021, with projections of prevalence to 2050. Lancet 402, 203–234 (2023). PMID 37356446.

21. GBD 2019 Hepatitis B Collaborators. Global, regional, and national burden of hepatitis B, 1990-2019. Lancet Gastroenterology & Hepatology 7, 796–829 (2022). PMID 35738290.

22. Bray, F. et al. Global cancer statistics 2022: GLOBOCAN estimates of incidence and mortality worldwide for 36 cancers in 185 countries. CA: A Cancer Journal for Clinicians 74, 229–263 (2024). PMID 38572751.

23. Alam, R. et al. Managing diagnostic uncertainty in primary care: a systematic critical review. BMC Family Practice 18, 79 (2017). PMID 28784088.

24. Jones, D. et al. Safety netting for primary care: evidence from a literature review. British Journal of General Practice 69, e70–e79 (2019). PMID 30510099.

25. Pauker, S. G. & Kassirer, J. P. The threshold approach to clinical decision making. New England Journal of Medicine 302, 1109–1117 (1980). PMID 7366635.

26. Eisenberg, J. M. & Hershey, J. C. Derived thresholds: determining the diagnostic probabilities at which clinicians initiate testing and treatment. Medical Decision Making 3, 155–168 (1983). PMID 6415358.

27. Kleindorfer, D. O. et al. 2021 guideline for the prevention of stroke in patients with stroke and transient ischemic attack. Stroke 52, e364–e467 (2021). PMID 34024117.

28. American Diabetes Association Professional Practice Committee. Pharmacologic approaches to glycemic treatment: Standards of Care in Diabetes 2026. Diabetes Care 49, S181–S206 (2026). PMID 41358900.

29. American Diabetes Association Professional Practice Committee. Chronic kidney disease and risk management: Standards of Care in Diabetes 2026. Diabetes Care 49, S219–S236 (2026). PMID 41358881.

30. European Association for the Study of the Liver. EASL Clinical Practice Guidelines on the management of hepatitis B virus infection. Journal of Hepatology 82, 123–160 (2025). PMID 40348683.

31. World Health Organization. Guidelines for the prevention, diagnosis, care and treatment for people with chronic hepatitis B infection. WHO guideline (2024). PMID 40424433.

32. Riely, G. J. et al. Non-Small Cell Lung Cancer, Version 4.2024, NCCN Clinical Practice Guidelines in Oncology. Journal of the National Comprehensive Cancer Network 22, 249–274 (2024). PMID 38754467.

33. Bazhenova, L. et al. Therapy for Stage IV Non-Small Cell Lung Cancer With Driver Alterations: ASCO Living Guideline, Version 2024.2. Journal of Clinical Oncology 42, 4321–4337 (2024). PMID 39531596.

34. Reuss, J. E. et al. Therapy for Stage IV Non-Small Cell Lung Cancer Without Driver Alterations: ASCO Living Guideline, 2026.3.0. Journal of Clinical Oncology 44, 1012–1031 (2026). PMID 41632927.

35. Liu, L. et al. Using natural language processing to extract information from clinical text in electronic medical records for populating clinical registries: a systematic review. Journal of the American Medical Informatics Association 33, 144–158 (2026). PMID 41093296.

36. Uzuner, O. et al. 2010 i2b2/VA challenge on concepts, assertions, and relations in clinical text. Journal of the American Medical Informatics Association 18, 552–556 (2011). PMID 21685143.

37. Rink, B., Harabagiu, S. & Roberts, K. Automatic extraction of relations between medical concepts in clinical texts. Journal of the American Medical Informatics Association 18, 594–600 (2011). PMID 21846787.

38. Sun, W., Rumshisky, A. & Uzuner, O. Evaluating temporal relations in clinical text: 2012 i2b2 Challenge. Journal of the American Medical Informatics Association 20, 806–813 (2013). PMID 23564629.

39. Chen, X. et al. Generating synthetic electronic health record data: a methodological scoping review with benchmarking on phenotype data and open-source software. Journal of the American Medical Informatics Association 32, 998–1012 (2025). PMID 40460023.

40. Davis, S. E. et al. A framework for understanding label leakage in machine learning for health care. Journal of the American Medical Informatics Association 30, 1444–1451 (2023). PMID 37669138.

41. Uzuner, O. et al. Community annotation experiment for ground truth generation for the i2b2 medication challenge. Journal of the American Medical Informatics Association 17, 519–523 (2010). PMID 20819855.

42. Chen, Y. et al. A study of active learning methods for named entity recognition in clinical text. Journal of Biomedical Informatics 58, 11–18 (2015). PMID 26385377.

43. Fu, S. et al. A taxonomy for advancing systematic error analysis in multi-site electronic health recordbased clinical concept extraction. Journal of the American Medical Informatics Association 31, 1607–1618 (2024). PMID 38742455.

44. Weiskopf, N. G. et al. Biases in electronic health record data for generating real-world evidence: an overview. Journal of the American Medical Informatics Association 31, 134–145 (2024). PMID 38273982.

45. Yang, R. et al. A roadmap for medical large language models. Medical Review 4, 130–156 (2026). PMID 42388248.

46. Zhang, A. et al. Shifting machine learning for healthcare from development to deployment and from models to data. Nature Biomedical Engineering 6, 1330–1345 (2022). PMID 35788685.

47. Goddard, K., Roudsari, A. & Wyatt, J. C. Automation bias: a systematic review of frequency, effect mediators, and mitigators. Journal of the American Medical Informatics Association 19, 121–127 (2012). PMID 21685142.

48. Vasey, B. et al. Association of clinician diagnostic performance with machine learning-based decision support systems: a systematic review. JAMA Network Open 4, e211276 (2021). PMID 33704476.

49. Tordjman, M. et al. Comparative benchmarking of the DeepSeek large language model on medical tasks and clinical reasoning. Nature Medicine 31, 1912–1922 (2025). PMID 40267969.

50. Goh, E. et al. Large language model influence on diagnostic reasoning: a randomized clinical trial. JAMA Network Open 7, e2440969 (2024). PMID 39466245.

51. Clusmann, J. et al. Safety and security of large language models in healthcare. Nature 650, 24–36 (2026). PMID 42618758.

52. Vasey, B. et al. Reporting guideline for the early-stage clinical evaluation of decision support systems driven by artificial intelligence: DECIDE-AI. Nature Medicine 28, 924–933 (2022). PMID 35585198.

53. Liu, X. et al. Reporting guidelines for clinical trial reports for interventions involving artificial intelligence: the CONSORT-AI extension. Nature Medicine 26, 1364–1374 (2020). PMID 32908283.

54. Rivera, S. C. et al. Guidelines for clinical trial protocols for interventions involving artificial intelligence: the SPIRIT-AI extension. Nature Medicine 26, 1351–1363 (2020). PMID 32908284.

55. Collins, G. S. et al. TRIPOD+AI statement: updated guidance for reporting clinical prediction models that use regression or machine learning methods. BMJ 385, e078378 (2024). PMID 38626948.

56. Moons, K. G. M. et al. PROBAST+AI: an updated quality, risk of bias, and applicability assessment tool for prediction models using regression or artificial intelligence methods. BMJ 388, e078609 (2025). PMID 40127903.

57. Norgeot, B. et al. Minimum information about clinical artificial intelligence modeling: the MI-CLAIM checklist. Nature Medicine 26, 1320–1324 (2020). PMID 32908275.

58. Miao, B. Y. et al. The MI-CLAIM-GEN checklist for generative artificial intelligence in health. Nature Medicine 31, 894–902 (2025). PMID 39915678.

